# Avoiding false positive SARS-CoV-2 rapid antigen test results with point-of-care molecular testing on residual test buffer

**DOI:** 10.1101/2022.02.18.22271189

**Authors:** Jason J LeBlanc, Gregory R. McCracken, Barbara Goodall, Todd F Hatchette, Lisa Barrett, John Ross, Ross J Davidson, Glenn Patriquin

## Abstract

**Objectives:** Antigen-based rapid diagnostic tests (Ag-RDTs) have been widely used for the detection of SARS-CoV-2 during the Covid-19 pandemic. In settings of low disease prevalence, such as asymptomatic community testing, national guidelines recommend molecular confirmation of positive Ag-RDT results. This often requires patients to be recalled for repeat specimen recollection and subsequent testing in reference laboratories. This project assessed the use of a point-of-care molecular method for SARS-CoV-2 detection on-site at a volunteer-led asymptomatic community testing site, using the residual test buffer (RTB) from positive Ag-RDTs.

**Methods:** The Abbott COVID-19 ID NOW assay was performed on RTB from two Ag-RDTs: the Abbott Panbio COVID-19 Ag Rapid Test Device and the BTNX Rapid Response COVID-19 Antigen Rapid Test Device. All RTBs were tested using real-time RT-PCR at a reference laboratory using the ThermoFisher TaqPath COVID-19 Combo kit which was used to assign positive Ag-RDTs results as true or false positives. Analytical specificity of the ID NOW was assessed with a panel of various respiratory organisms.

**Results:** Of 419 positive Ag-RDTs from 5148 tests performed, ID NOW testing of the RTB was positive in 100% of the samples characterized as true positives by RT-PCR. No SARS-CoV-2 detections by ID NOW were observed from 10 specimens characterized as false positive Ag-RDTs, or from contrived specimens with various respiratory organisms.

**Conclusions:** The use of on-site molecular testing on RTB provides a suitable option for rapid confirmatory testing of positive Ag-RDTs, thereby obviating the need for specimen recollection for molecular testing at local reference laboratories.

## Introduction

With their simplicity, speed, and scalability, antigen-based rapid diagnostic tests (Ag-RDTs) have been deployed worldwide to facilitate SARS-CoV-2 detection.^1-5^ Ag-RDT positive results has been associated with the ability to culture SARS-CoV-2 *in vitro* or viral loads consistent with transmissible virus; therefore, Ag-RDTs has been used as a surrogate for SARS-CoV-2 communicability.^6-10^ Nova Scotia was the first Canadian province to implement Ag-RDTs for self-perceived asymptomatic individuals in low-barrier volunteer-led community testing centres, to identify individuals at high risk of transmitting SARS-CoV-2 that might otherwise have gone unnoticed.^11,12^ Following national guidelines, individuals with positive Ag-RDTs were asked to return to testing centres for specimen recollection, and confirmatory testing using nucleic acid amplification tests (NAATs) performed at local reference laboratories. To streamline confirmation of positive Ag-RDTs, direct NAAT testing on the Ag-RDT residual test buffer (RTB) was evaluated.

Like others^13^, our previous study demonstrated high sensitivity for SARS-CoV-2 detection using RTB from nasopharyngeal and nasal swab collections.^11^ RTB obviated the need for specimen recollection for NAAT-based confirmation of Ag-RDT results, but RTB processing remained at the reference laboratory. To further optimize community testing strategies, this project evaluated a portable NAAT-based rapid diagnostic test (NAAT-RDT) for on-site confirmation of Ag-RDTs-positive results at the community testing centers. The COVID-19 assay on the Abbott ID NOW instrument is an NAAT-RDT that uses isothermal technology that is amenable to point-of-care applications.^4^ This NAAT-RDT is simple and provides rapid results with high sensitivity and specificity, but its single-specimen processing limits its scalability for testing large populations.^14-16^ Instead, this NAAT-RDT was evaluated for rapid confirmation of SARS-CoV-2 using RTB from positive Ag-RDTs, at the site of sample collection (Table 1).

**Table 1.** Summary of ID NOW results from all study phases.

| Category |  |  | ID NOW results* |  |  |  |
| --- | --- | --- | --- | --- | --- | --- |
|  |  |  | Nasal<br>(n=164) | Throat<br>(n=93) | Combined nasal/throat<br>(n=162) | Total<br>(n=419) |
| Antigen status | Ag+/NAAT+ |  | 100.0% (132/132) | 100.0% (66/66) | 100% (156/156) | 100.0% (354/354) |
|  | Ag+/NAAT- |  | 0.0% (0/4) | N/A | 0.0% (0/6) | 0.0% (0/10) |
|  | Ag-/NAAT+ |  | 82.1% (23/28) | 92.6% (25/27) | N/A | 87.3% (48/55) |
| Antigen score | Ag+/NAAT+ | 3+ | 100.0% (32/32) | 100.0% (13/13) | 100.0% (41/41) | 100.0% (86/86) |
|  |  | 2+ | 100% (43/43) | 100.0% (22/22) | 100.0% (52/52) | 100.0% (117/117) |
|  |  | 1+ | 100.0% (36/36) | 100.0% (19/19) | 100.0% (33/33) | 100.0% (88/88) |
|  |  | +/- | 100.0% (21/21) | 100.0% (12/12) | 100.0% (30/30) | 100.0% (63/63) |
|  | Ag+/NAAT- | 1+ | 0.0% (0/1) | N/A | 0.0% (0/2) | 0.0% (0/3) |
|  |  | +/- | 0.0% (0/3) | N/A | 0.0% (0/4) | 0.0% (0/7) |
| Ct value** | Ag+/NAAT+ | <25 | 100.0% (28/28) | 100.0% (6/6) | 100.0% (62/62) | 100.0% (96/96) |
|  |  | 25 to <30 | 100.0% (58/58) | 100.0% (30/30) | 100.0% (68/68) | 100.0% (156/156) |
|  |  | ≥30 | 100.0% (46/46) | 100.0% (30/30) | 100.0% (26/26) | 100.0% (102/102) |
|  | Ag-/NAAT+ | 25 to <30 | N/A | 100.0% (4/4) | N/A | 100.0% (4/4) |
|  |  | ≥30 | 82.1% (23/28) | 91.3% (21/23) | N/A | 86.3% (44/51) |

\*ID Now for individual test and project phases are provided in Tables S1, S2, and S3. \*\*Ct values were categorized based on the N gene of the TaqPath real-time RT-PCR. Abbreviations: antigen (Ag); antigen-based rapid diagnostic test (Ag-RDT); threshold cycle (Ct); nucleic acid amplification test (NAAT); residual test buffer (RTB).

The assessment was performed in two stages on asymptomatic individuals presenting to urban rapid testing sites. The first overlapping with the ISNOT project (designed to compare SARS-CoV-2 Ag-RDT results from self-administered nasal and throat collections)^12^ (Table S1 and S2), and the second was an extension of the project where additional positive Ag-RDT RTBs were tested over a subsequent two-week period (Table S3). In both cases, Ag-RDT self-testing using the Panbio COVID-19 Ag Rapid Test Device (Abbott Rapid Diagnostics, Jena, Germany) or the Rapid Response COVID-19 Antigen Rapid Test Device (BTNX Inc., Markham, ON) was used. Ag-RDTs were interpreted according to manufacturer instructions, and SARS-CoV-2 target bands were graded with scores of 0 (negative), +/- (barely visible), or 1+, 2+ or 3+ relative to the intensity of the control band. RTB from either Ag-RDT were subjected to two NAATs. The COVID-19 ID Now assay (Abbott Diagnostics, Scarborough, MA) was performed on-site following manufacturer instructions, except two drops of RTB that were added to the sample chamber prior to processing the original Ag-RDT collection swab. The remaining RTB and swab were transported to a central laboratory in the Ag-RDT reaction tube, and 200 µl of viral transport medial (VTM) (Rodoxica, Little Rock, AR) was added to the tube. Following vortexing for 10 seconds, 200 µl of VTM/RTB fluid was subjected to a total nucleic acid extraction (TNA) on a MagNA Pure 96 or LC 2.0 instrument (Roche Diagnostics ltd., Roltkreuz, Switzerland), and 5 µl of the 50 µl of eluted TNAs were used as template for real-time RT-PCR using the TaqPath COVID-19 Combo Kit (Life Technologies Corp., Frederick, MD).

Of 1472 individuals who consented to the ISNOT project^12^, 159 and 58 were positive using the Panbio (Table 1) and BTNX (Table S1) Ag-RDTs, respectively. The ID NOW was positive for all Ag-RDT positive RTB samples, regardless of anatomical site of collection, Ag-RDT method used, antigen score, or RT-PCR threshold cycle (Ct) value (Tables S1 and S2). No false positives were identified, suggesting high specificity. This data supports use of the NAAT-RDT to quickly rule-in SARS-CoV-2 using RTB from positive Ag-RDTs, thereby ruling out false positive Ag-RDT reactions. However, further testing was needed to verify if false positive Ag-RDTs would be negative with the ID NOW assay. First, highly concentrated nucleic acids from various respiratory microorganisms were spiked into 300 µl Panbio buffer and tested with the ID NOW assay. The assay detected a variety of SARS-CoV-2 lineages, but no cross-reactions were observed with other respiratory organisms (Table S4). Then, for two weeks following the ISNOT project, RTB from positive Ag-RDT reactions were subjected to ID NOW and RT-PCR testing (Table S3). Of 3676 individuals tested, 147 had positive Ag-RDTs, and 137 were positive for both ID NOW and RT-PCR. There were 10 false positive Ag-RDTs compared to RT-PCR, and these were also negative by ID NOW. Consistent with our previous study^11^, false positive Ag-RDTs were described as having barely visible target bands, with antigen scores +/- or 1+. The proportion of false positives observed (10/3676 or 0.3%) is consistent with manufacturer and literature claims, where false positive reactions are rare at approximately 0.4%^18,19^.

Specificity was the focus for the intended use of the ID NOW in this investigation. Sensitivity analyses would require IDNOW and RT-PCR testing on all Ag-RDT RTB negative specimens, which would not typically be performed in community-based surveillance. Some sensitivity data was captured during the ISNOT quality initiative^12^, as the ID NOW was performed in parallel on Ag-RDT RTB from paired swabs samples from positive individuals. As such, some specimens were negative by Ag-RDT for one swab type of the paired collection, but positive results were obtained by ID NOW and/or RT-PCR. While Ag-RDTs appear less sensitive than NAATs (as seen in Tables S1 and S2), it has been argued that the additional detections by NAATs often represent remnant RNA from resolved infections, when the risk for transmission is low.^20-23^ Alternatively, it may represent periods of early infection that are short-lived in population-based testing and can be mitigated by frequent testing over time with Ag-RDTs.^20-23^ Importantly, ID NOW confirmed all true positive Ag-RDTs, as well as detected 88.2% (30/34) and 85.7% (18/21) of negative Panbio and BTNX Ag-RDT RTBs that tested positive by RT-PCR, respectively (Tables S1 and S2). Discrepant results between ID NOW and RT-PCR were in specimens with Ct values ≥30, suggesting low viral loads. Altogether, the ID NOW was found to be sufficiently sensitive to be used as a confirmatory method for Ag-RDTs.

Overall, ID NOW confirmed 409 true positive Ag-RDT results, ruled out 10 false positives, and there was no cross-reactivity with other respiratory organisms. The applications of rapid NAAT-RDT confirmation of positive Ag-RDTs on-site using RTB obviates the need for individuals to return for repeat specimen collection for NAAT testing at local reference laboratories, as well as the need for trained personnel for shipping biological samples to reference laboratories. With recent surges of SARS-CoV-2 activity with the highly transmissible Omicron variant, many clinical laboratories were overwhelmed with high testing demands, hampering their ability to support confirmation for Ag-RDT-positive results. Given wide community spread, and the low proportion of false positive results during this period of high disease prevalence, NAAT-based confirmation of Ag-RDT results was not prioritized. However, in the wake of pandemic waves as disease prevalence decreases, the possibility of false positive Ag-RDT increases and confirmatory testing for Ag-RDT will again become important to consider.^24,25^ The use of RTB testing with NAAT-RDTs provides a feasible and accurate option for rapid confirmatory testing of positive Ag-RDTs at community testing sites.

## Data Availability

All data produced in the present work are contained in the manuscript

## Acknowledgements

The authors are indebted to the community participants for this evaluation, and to the many Test-to-Protect volunteers and coaches who have worked countless hours throughout the pandemic to ensure the safety of their community and community members. The authors would also like to thank the clinical laboratories across Nova Scotia, who have supported confirmation of Ag-RDTs results throughout the pandemic.

## Ethics

This project was deemed a quality initiative and was therefore exempt from review by the Nova Scotia Health Research Ethics Board (submission number 1027644). Specimens tested were obtained from consenting participants, and all data related were provided anonymized, de-identified, and were used solely with the intent to evaluate the performance characteristics of the different swab types for rapid antigen testing programs used in Nova Scotia.

## Author contributions

All authors were involved in the design, data acquisition, and data interpretation. JL, GP, and TH drafted the initial manuscript, with all authors contributing, and agree with the content of the final version.

