## Supplemental Table S1 for "Avoiding false positive SARS-CoV-2 rapid antigen test results with point-of-care molecular testing on residual test buffer"

**Table S1.** ID NOW results from the RTB of the Panbio Ag-RDT during the ISNOT project.

| **Category** | | | **ID NOW results** | | | |
| --- | --- | --- | --- | --- | --- | --- |
|  |  |  | **Nasal**  **(n=96)** | **Throat**  **(n=55)** | **Combined nasal/throat**  **(n=42)** | **Total**  **(n=193)** |
| **Antigen status** | **Ag+/NAAT+** | | 100.0% (77/77) | 100.0% (40/40) | 100.0% (42/42) | 100.0% (159/159) |
|  | **Ag+/NAAT-** | | N/A | N/A | N/A | N/A |
|  | **Ag-/NAAT+** | | 78.9% (15/19) | 100.0% (15/15) | N/A | 88.2% (30/34) |
| **Antigen score** | **Ag+/NAAT+** | **3+** | 100.0% (26/26) | 100.0% (13/13) | 100.0% (13/13) | 100.0% (52/52) |
|  |  | **2+** | 100.0% (23/23) | 100.0% (10/10) | 100.0% (16/16) | 100.0% (49/49) |
|  |  | **1+** | 100.0% (20/20) | 100.0% (11/11) | 100.0% (8/8) | 100.0% (39/39) |
|  |  | **+/-** | 100.0% (8/8) | 100.0% (6/6) | 100.0% (5/5) | 100.0% (19/19) |
| **Ct value*** | **Ag+/NAAT+** | **<25** | 100.0% (16/16) | 100.0% (2/2) | 100.0% (8/8) | 100.0% (26/26) |
|  |  | **25 to <30** | 100.0% (31/31) | 100.0% (18/18) | 100.0% (22/22) | 100.0% (71/71) |
|  |  | **≥30** | 100.0% (30/30) | 100.0% (20/20) | 100.0% (12/12) | 100.0% (62/62) |
|  | **Ag-/NAAT+** | **<25** | N/A | N/A | N/A | N/A |
|  |  | **25 to <30** | N/A | N/A | N/A | N/A |
|  |  | **≥30** | 78.9% (15/19) | 100.0% (15/15) | N/A | 88.2% (30/34) |

*Ct values were categorized based on the N gene of the TaqPath real-time RT-PCR. Abbreviations: antigen (Ag); antigen-based rapid diagnostic test (Ag-RDT); threshold cycle (Ct); nucleic acid amplification test (NAAT); residual test buffer (RTB).
