## Supplemental Table S2 for "Avoiding false positive SARS-CoV-2 rapid antigen test results with point-of-care molecular testing on residual test buffer"

**Table S2.** ID NOW results from the RTB of the BTNX Ag-RDT during the ISNOT project.

| **Category** | | | **IDNOW results** | | |
| --- | --- | --- | --- | --- | --- |
|  |  |  | **Nasal**  **(n=41)** | **Throat**  **(n=38)** | **Total**  **(n=79)** |
| **Antigen status** | **Ag+/NAAT+** | | 100.0% (32/32) | 100.0% (26/26) | 100.0% (58/58) |
|  | **Ag+/NAAT-** | | N/A | N/A | N/A |
|  | **Ag-/NAAT+** | | 88.9% (8/9) | 83.3% (10/12) | 85.7% (18/21) |
| **Antigen score** | **Ag+/NAAT+** | **2+** | 100.0% (16/16) | 100.0%(12/12) | 100.0% (28/28) |
|  |  | **1+** | 100.0% (12/12) | 100.0% (8/8) | 100.0% (20/20) |
|  |  | **+/-** | 100.0% (4/4) | 100.0% (6/6) | 100.0% (10/10) |
| **Ct value*** | **Ag+/NAAT+** | **<25** | 100.0% (1/1) | 100.0% (4/4) | 100.0% (5/5) |
|  |  | **25 to <30** | 100.0% (20/20) | 100.0% (12/12) | 100.0% (32/32) |
|  |  | **≥30** | 100.0% (11/11) | 100.0% (10/10) | 100.0% (21/21) |
|  | **Ag-/NAAT+** | **<25** | N/A | N/A | N/A |
|  |  | **25 to <30** | N/A | 100.0% (4/4)^**^ | 100.0% (4/4)** |
|  |  | **≥30** | 88.9% (8/9) | 75.0% (6/8) | 82.4% (14/17) |

*Ct values were categorized based on the N gene of the TaqPath real-time RT-PCR. **Ct values falling into this category were 27.43, 27.71, 29.63, and 29.87. Abbreviations: antigen (Ag); antigen-based rapid diagnostic test (Ag-RDT); threshold cycle (Ct); nucleic acid amplification test (NAAT); residual test buffer (RTB).
