## Supplemental Table S3 for "Avoiding false positive SARS-CoV-2 rapid antigen test results with point-of-care molecular testing on residual test buffer"

**Table S3.** Summary of the supplemental testing to identify false positive with the Panbio Ag-RDT.

| **Category** | | | **ID NOW results** | | |
| --- | --- | --- | --- | --- | --- |
|  |  |  | **Nasal**  **(n=27)** | **Combined nasal/throat**  **(n=120)** | **Total**  **(n=147)** |
| **Antigen status** | **Ag+/NAAT+** | | 100.0% (23/23) | 100.0% (114/114) | 100.0% (137/137) |
|  | **Ag+/NAAT-** | | 0.0% (0/4) | 0.0% (0/6) | 0.0% (0/10) |
|  | **Ag-/NAAT+** | | N/A | N/A | N/A |
| **Antigen score** | **Ag+/NAAT+** | **3+** | 100.0% (6/6) | 100.0% (28/28) | 100.0% (34/34) |
|  |  | **2+** | 100.0% (4/4) | 100.0% (36/36) | 100.0% (40/40) |
|  |  | **1+** | 100.0% (4/4) | 100.0% (25/25) | 100.0% (29/29) |
|  |  | **+/-** | 100.0% (9/9) | 100.0% (25/25) | 100.0% (34/34) |
|  | **Ag+/NAAT-** | **3+** | N/A | N/A | N/A |
|  |  | **2+** | N/A | N/A | N/A |
|  |  | **1+** | 0.0% (0/1) | 0.0% (0/2) | 0.0% (0/3) |
|  |  | **+/-** | 0.0% (0/3) | 0.0% (0/4) | 0.0% (0/7) |
| **Ct value*** | **Ag+/NAAT+** | **<25** | 100.0% (11/11) | 100.0% (54/54) | 100.0% (65/65) |
|  |  | **25 to <30** | 100.0% (7/7) | 100.0% (46/46) | 100.0% (53/53) |
|  |  | **≥30** | 100.0% (5/5) | 100.0% (14/14) | 100.0% (19/19) |

*Ct values were categorized based on the N gene of the TaqPath real-time RT-PCR. Abbreviations: antigen (Ag); antigen-based rapid diagnostic test (Ag-RDT); threshold cycle (Ct); nucleic acid amplification test (NAAT); residual test buffer (RTB).
