## Supplemental Table S4 for "Avoiding false positive SARS-CoV-2 rapid antigen test results with point-of-care molecular testing on residual test buffer"

**Table S4.** Analytical specificity panel for evaluation of the ID NOW COVID-19 assay.

| **Microorganism** | | **ID NOW result** |
| --- | --- | --- |
| **SARS-CoV-2 lineages** | |  |
|  | SARS-CoV-2 (lineage A.1) | POS |
|  | SARS-CoV-2 (lineage B.1.438) | POS |
|  | SARS-CoV-2 (lineage B.1.1.157) | POS |
|  | SARS-CoV-2, alpha (lineage B.1.1.7) | POS |
|  | SARS-CoV-2, beta (lineage B.1.351) | POS |
|  | SARS-CoV-2, gamma (lineage P.1) | POS |
|  | SARS-CoV-2, delta (lineage B.1.617.2) | POS |
|  | SARS-CoV-2, epsilon (lineage B.1.427) | POS |
|  | SARS-CoV-2, iota (lineage B.1.1.526) | POS |
|  | SARS-CoV-2, lambda (lineage C.37) | POS |
|  | SARS-CoV-2, omicron (lineage B.1.1.529) | POS |
| **Other human coronaviruses (hCoV)** | |  |
|  | SARS-CoV-1 | NEG |
|  | MERS-CoV | NEG |
|  | hCoV, 229E | NEG |
|  | hCoV, OC43 | NEG |
|  | hCoV, NL63 | NEG |
|  | hCoV, HKU1 | NEG |
| **Other respiratory viruses** | |  |
|  | Influenza A virus (FluA), subtype H3N2 | NEG |
|  | Influenza A virus (FluA), subtype H3N2 | NEG |
|  | Influenza B virus (FluB), Yamagata lineage | NEG |
|  | Influenza B virus (FluB), Victoria lineage | NEG |
|  | Respiratory syncytial virus (RSV-A), type A | NEG |
|  | Respiratory syncytial virus (RSV-B), type B | NEG |
|  | Parainfluenza virus, type 1 (P1V) | NEG |
|  | Parainfluenza virus, type 2 (P2V) | NEG |
|  | Parainfluenza virus, type 3 (P3V) | NEG |
|  | Parainfluenza virus, type 4 (P4V) | NEG |
|  | Human metapneumovirus (hMPV) | NEG |
|  | Human adenovirus (hAdV), serogroup A | NEG |
|  | Human adenovirus (hAdV), serogroup B | NEG |
|  | Human adenovirus (hAdV), serogroup C | NEG |
|  | Human adenovirus (hAdV), serogroup D | NEG |
|  | Human adenovirus (hAdV), serogroup E | NEG |
|  | Human adenovirus (hAdV), serogroup F | NEG |
|  | Human rhinovirus, A (HRV-A) | NEG |
|  | Human rhinovirus, B (HRV-B) | NEG |
|  | Human enterovirus (HEV), echo9 | NEG |
|  | Human enterovirus (HEV), coxsackievirus A | NEG |
|  | Human enterovirus (HEV), paraechovirus | NEG |
|  | Human enterovirus (HEV), enterovirus D-68 | NEG |
|  | Human bocavirus (HBoV) | NEG |
| **Other viruses** | |  |
|  | Measles virus, genotype A | NEG |
|  | Measles virus, genotype B3 | NEG |
|  | Measles virus, genotype D8 | NEG |
|  | Mumps virus, genotype A | NEG |
|  | Mumps virus, genotype C | NEG |
|  | Mumps virus, genotype G | NEG |
|  | Rubella virus, genotype 1B-F | NEG |
|  | Rubella virus, genotype 2B | NEG |
|  | Human immunodeficiency virus 1(HIV-1) | NEG |
|  | Hepatitis C virus (HCV), genotype 3 | NEG |
|  | Hepatitis B virus (HBV) | NEG |
|  | Herpes simplex virus 1 (HSV-1) [human herpesvirus 1 (HHV-1)] | NEG |
|  | Herpes simplex virus 2 (HSV-2) [human herpesvirus 2 (HHV-2)] | NEG |
|  | Varicella zoster virus (VZV) [human herpesvirus 3 (HHV-3)] | NEG |
|  | Epstein Barr virus (EBV) [human herpesvirus 4 (HHV-4)] | NEG |
|  | Cytomegalovirus (CMV) [human herpesvirus 4 (HHV-4)] | NEG |
| **Other microorganisms** | |  |
|  | *Chlamydia pneumoniae* | NEG |
|  | *Haemophilus influenzae* | NEG |
|  | *Legionella pneumophila* | NEG |
|  | *Mycoplasma pneumoniae* | NEG |
|  | *Bordetella pertussis* | NEG |
|  | *Streptococcus pneumoniae* | NEG |
|  | *Streptococcus oralis* | NEG |
|  | *Streptococcus salivarius* | NEG |
|  | *Staphylococcus epidermis* | NEG |
|  | *Staphylococcus aureus* | NEG |
|  | *Pseudomonas aeruginosa* | NEG |
|  | *Mycobacterium tuberculosis* | NEG |
|  | *Pneumocystis jirovecii (PJP)* | NEG |
|  | *Candida albicans* | NEG |
|  | *Corynebacterium* sp. | NEG |
